# mTICI 1 vs mTICI 0 before endovascular stroke treatment in middle cerebral artery M1-occlusion – predictor for revascularization success and outcome?

**DOI:** 10.1101/2022.12.21.22283688

**Authors:** Jessica Jesser, Charlotte S. Weyland, Arne Potreck, Ulf Neuberger, Michael O. Breckwoldt, Min Chen, Silvia Schönenberger, Martin Bendszus, Markus A. Möhlenbruch

## Abstract

**Background:** Endovascular stroke treatment (EST) has become the treatment of choice for middle cerebral artery (MCA) M1-segment occlusions. Little is known about the implications for revascularization success of occlusions with persisting antegrade perfusion before initiation of treatment (modified Treatment In Cerebral Ischemia (mTICI 1)) compared to a complete occlusion (mTICI 0). Here, we compared the impact of these two states of target vessel occlusion on recanalization success and clinical outcome.

**Methods:** Retrospective, single-center analysis of patients treated for M1-segment MCA occlusion with EST from 01/2015 until 05/2020 in a tertiary stroke center. Primary study endpoint was successful recanalization (mTICI 2c-3) after one thrombectomy attempt (*first pass effect*). Secondary endpoints were the clinical outcome (as modified Rankin Scale 90 days after stroke onset) and the complication rate. The two study groups were compared in univariate analysis including patient characteristics and procedural details.

**Results:** In this study, 422/581 patients (72.6 %) presented with complete M1-occlusion compared to 159/581 (27.4 %) with incomplete M1-occlusion. Neither did the rate of FPE differ between the study groups nor the rate of procedural complications (mTICI 0: 10 (2.4%), mTICI 1: 1 (0.6%), p = 0.304). Patients with incomplete initial occlusion showed a lower mRS at discharge (median (IQR) mTICI0: 4 (3-5) vs. mTICI1: 3 (2 – 6), p = 0.014), but a comparable mRS 90 days after stroke onset (mTICI0: 3 (2-6) vs. mTICI:1 4 (2-6), p = 0.479).

**Conclusion:** Complete M1-occlusions (mTICI 0) and incomplete occlusions (mTICI 1) show the same recanalization success and complication rate as well as a comparable clinical outcome. Thus, incomplete M1-occlusions should be treated with the same urgency as initial complete occlusions.

## Introduction

Since endovascular stroke treatment (EST) became a standard treatment option in acute ischemic stroke in recent years^1^, many procedural and technical details were uncovered that are associated with a higher chance of clinical recovery after ischemic stroke and EST. Among these, successful revascularization of the target vessel occlusion (TVO) plays a pivotal role for a favorable clinical outcome ^2^.

So far, different types of angiographic occlusion shapes were described predicting the ensuing revascularization success. Baek et al. found a difference between truncal-type and branching type occlusion with more stent-retriever failure for truncal-type occlusions^3^. Consoli et al. reported differences of regular and irregular occlusion shape in responding to stent-retriever thrombectomy and contact aspiration^4^. Further occlusion shapes described so far are the so-called meniscus sign and the claw sign, referring to angiographically different shapes of TVO before EST. Miranda et al. found the meniscus sign not to be associated with good clinical outcome or revascularization success^5^, while Yamamoto et al. report that the claw sign might predict revascularization success^6^.

All of the described occlusion types share that the target vessel is completely occluded. According to the modified Treatment in Cerebral Ischemia (mTICI) score, the antegrade contrast flow is completely arrested without further perfusion (mTICI 0). In contrast, little is known about TVOs with persisting minimal antegrade perfusion past the initial occlusion but limited distal branch filling with little or slow distal perfusion (mTICI 1). The aim of this study was to identify differences in the revascularization success comparing mTICI 0 and mTICI 1 occlusions in middle cerebral artery M1-segment occlusions (figure 1). These two scenarios where compared using a well-established predictor of the clinical outcome and valid parameter to compare thrombectomy scenarios, techniques and material: the so called “*first pass effect*” (FPE)^7^. FPE refers to a complete target vessel recanalization with one thrombectomy attempt^8^. Furthermore, the applied thrombectomy techniques, contact aspiration or stent-retriever thrombectomy, and patient characteristics were compared between the two study groups. The clinical outcome of the two study groups were compared for the short-term follow up of modified Rankin Scale (mRS) at discharge as well as long-term outcome as mRS 90 days after stroke onset.

**Figure 1.**
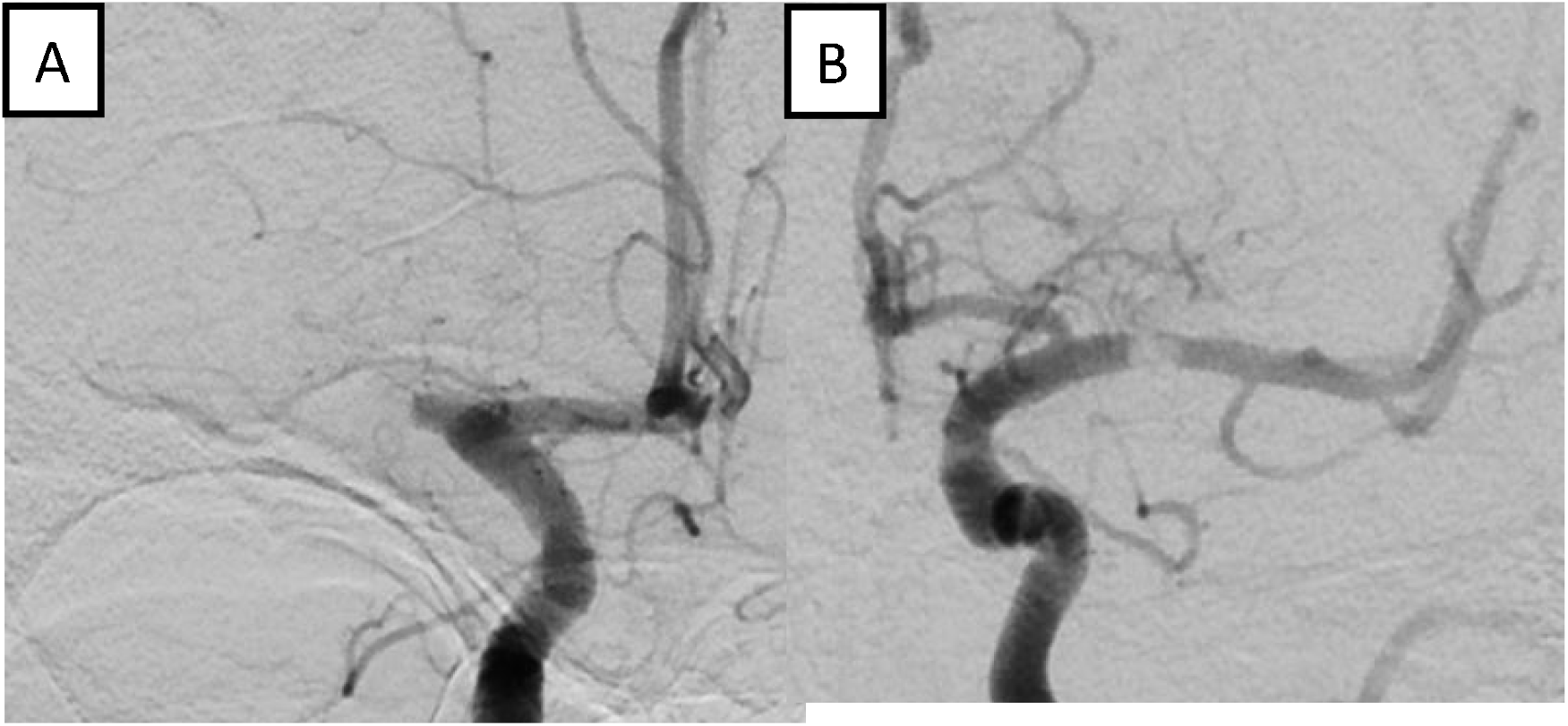
**A**: right middle cerebral artery (MCA) M1-occlusion (modified Treatment In Cerebral Ischemia (mTICI) 0), **B:** left MCA M1-occlusion (mTICI 1)

## Methods

This is a retrospective single-center analysis with prospectively collected data from a tertiary stroke care center. Patients were included with a middle cerebral artery M1-occlusion who were eligible for endovascular stroke treatment between 01/2015 and 05/2020. The ethics committee *(blinded for peer review)* approved this work. Patient consent was waived due to the retrospective nature of this study. Patients with a complete occlusion of the extracranial internal carotid artery (tandem occlusion) were excluded because of the missing imaging of the intracranial TVO before intervention. Also, patients with poor quality of initial angiographic imaging of the TVO, e.g. due to motion artifacts and patients with no thrombectomy pass performed, were excluded – see figure 2.

**Figure 2.**
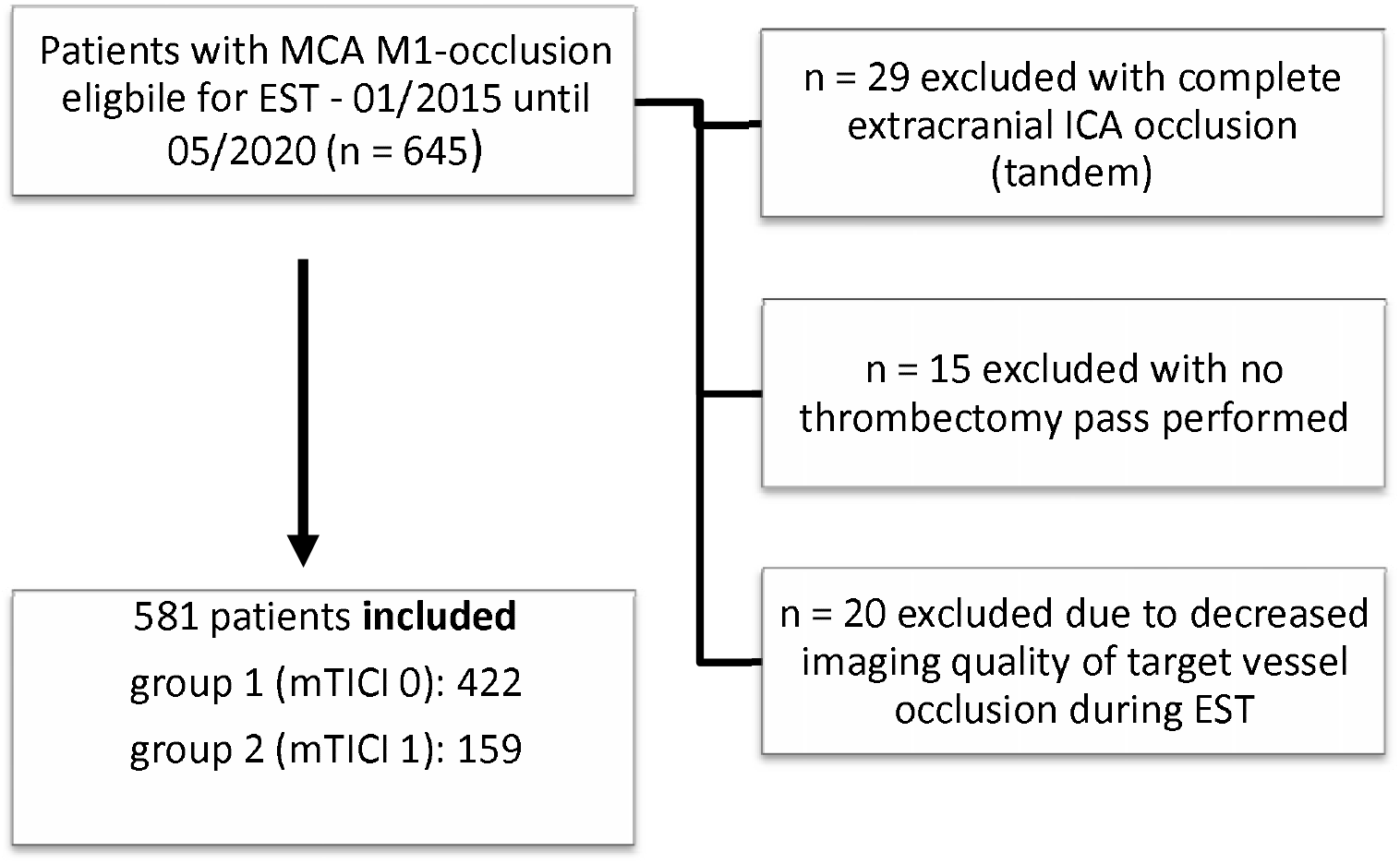
Flow chart of patient selection and study groups. EST = endovascular stroke treatment, MCA – middle cerebral artery, mTICI – modified Treatment in Cerebral Ischemia

### Study groups and study endpoints

The two study groups were defined as group 1 with complete occlusion of the MCA M1-segment TVO (mTICI 0) and group 2 with persisting antegrade perfusion of the TVO (mTICI 1). The EST’s angiographic imaging was reviewed by two experienced interventional neuroradiologists blinded to the clinical outcome (with 6 and 12 years of clinical practice) to assess the mTICI score in initial imaging of the TVO before intracranial probing. Consensus reading was performed in case of differing assessments. The primary study endpoint was *first pass effect* (FPE) with complete reperfusion after one thrombectomy maneuver (stent-retriever thrombectomy or contact aspiration), defined as mTICI 2c or 3. Secondary endpoints were the occurrence of procedural complications (vessel perforation, dissection, vasospasm and thromboembolism during EST) and the clinical outcome at discharge and 90 days after stroke onset according to the modified Rankin Scale.

### EST treatment protocol and neurological assessment

In all patients, the choice of material and the primary thrombectomy approach (contact aspiration or stent-retriever thrombectomy under continuous aspiration) was decided by the interventionalist. In our center the standard set-up for EST in the anterior circulation is a triaxial system. This comprises a balloon-guided catheter (Merci 9F 95cm or Flowgate 8F 95 cm, Stryker, Kalamazoo, USA), an intermediate catheter (e.g. Sofia 5F/6F, Microvention, Aliso Viejo, California, USA) and a microcatheter/microwire system (mostly Rebar18 and Traxcess14). The two stent-retrievers being used the most in this study are Solitaire X (Medtronic, Irvin, CA) and Trevo (Stryker, Kalamazoo, USA). Stent-retriever thrombectomy attempts were performed during continuous aspiration using a VacLok-Syringe® at the aspiration catheter and a VacLok-Syringe at the guiding catheter. Conscious sedation or general anesthesia was used according to the patients clinical presentation. Anesthesia surveillance was performed by intensive care experienced neurologists. Clinical and neurological baseline characteristics (NIHSS and pre-stroke mRS) were assessed by neurologists. Clinical outcome after discharge (90 days after stroke onset) was performed in person or on telephone by neurologists or clinical scientific assistants.

### Data Acquisition and statistical analysis

Source data were generated from a prospectively collected stroke database. Additionally, all data included in the present analysis were validated retrospectively. Groups were compared for differences in thrombectomy technique, clinical outcome, imaging and procedural aspects with a Kruskal-Wallis test with Dunn’s correction for multiple comparisons or the Mann-Whitney U test when combining groups and comparing two groups only. Frequencies were compared with a Chi-Squared test for multiple groups or Fisher’s exact test for two groups in case groups were combined. Statistical analysis was conducted using SPSS 25.0. For all statistical tests the significance level was set to p = 0.05. Medians are given with interquartile range (IQR). All confidence intervals (CI) are quoted as 95% CI.

## Results

The patients included in the study analysis showed a complete target vessel occlusion in the majority of cases (mTICI 0: 72.6%; n = 422/581 patients) and a persisting antegrade perfusion (mTICI 1: 27.4 %; n = 159/581 patients). The patients presenting with complete occlusion (mTICI 0) showed a higher NIHSS compared to incomplete occlusions (NIHSS for mTICI 0, median (IQR): 16 (11 – 21) vs. mTICI 1 14 (8 – 19.5), p = 0.015). For patients with complete occlusion, the modified Rankin Scale (mRS) at discharge was higher (mRS median (IQR) in patients with mTICI 0: 4 (3 – 5) vs.mRS: 3 (2 – 6) in patients with mTICI 1, p = 0.014) despite a comparable pre-stroke mRS (in patients with mTICI 0, pre-stroke mRS was 1 (0 – 3) vs. pre-stroke mRS was 1 (0 – 2) in patients with mTICI 1, p = 0.6) while the patient age was higher in the mTICI 1 study group (mTICI 0: median age of 78 (67 – 82) vs. mTICI 1: median age of 79 (69 – 85), p = 0.043) – see also table 1. Pre-stroke anticoagulation or antiplatelet therapy was found more often in patients with incomplete initial TVO (mTICI0 33.9% vs. mTICI1 43.3%, p = 0.042). While comorbidities, procedure times (time from symptom onset to groin puncture and time from groin puncture to recanalization) and Alberta Stroke Program Early CT Score (ASPECTS) of baseline imaging were equally distributed between the study groups, patients with an incomplete occlusion showed a higher ASPECTS in follow-up imaging (mTICI 0: median ASPECTS of 7 (5 – 8) vs. mTICI 1: median ASPECTS of 8 (5.5-9), p = 0.045).

**Table 1.** patient characteristics and clinical outcome of study groups

|  | **mTICI 0 (n = 422)** | **mTICI 1 (n = 159)** | **p value** |
| --- | --- | --- | --- |
| **Male, n (%)** | 164 (38.9) | 70 (44.0) | 0.297 |
| **Wake-up, n (%)** | 142 (33.6) | 68 (42.8) | 0.078 |
| **Age, median (IQR)** | 78 (67 – 82) | 79 (69 – 85) | **0.043** |
| **Diabetes, n (%)** | 85 (20.1) | 32 (20.1) | 1.0 |
| **Hypertension, n (%)** | 287 (68) | 111 (69.8) | 0.907 |
| **Coronary artery disease, n (%)** | 82 (19.4) | 34 (21.4) | 0.725 |
| **Atrial fibrillation, n (%)** | 168 (39.8) | 70 (44.0) | 0.432 |
| **Antiplatelet therapy or Anticoagulation, n (%)** | 143 (33.9) | 69 (43.3) | **0.042** |
| **i.v. thrombolysis, n (%)** | 189 (44.8) | 51 (32.1) | **0.004** |
| **Baseline ASPECTS, median (IQR)** | 8 (7 – 10) | 9 (7 – 10) | 0.532 |
| **ASPECTS, follow up, median (IQR)** | 7 (5 – 8) | 8 (5.5 – 9) | **0.045** |
| **Extracranial ICA stenosis, n (%)** | 31 (7.3) | 6 (3.8) | 0.130 |
| **Time onset to groin puncture, median (IQR)** | 255 (166 – 430) | 275 (182 – 591) | 0.179 |
| **Time groin puncture to recanalization, median (IQR)** | 25 (17 – 36) | 24 (17 – 37) | 0.800 |
| **Baseline neurological presentation and clinical outcome in study groups** |  |  |  |
| **Baseline NIHSS, n (%)** | 16 (11 – 21) | 14 (8 – 19.5) | **0.015** |
| **NIHSS 24 hrs, median (IQR)** | 10.5 (4 – 19) | 8.5 (3 – 18) | 0.175 |
| **Pre-stroke mRS, median (IQR)** | 1 (0 – 3) | 1 (0 – 2) | 0.561 |
| mRS discharge, median (IQR) | 4 (3 – 5) | 3 (2 – 6) | **0.014** |
| mRS 90, median (IQR) | 3 (2 – 6) | 4 (2 – 6) | 0.479 |

Both study groups did not show any difference in the recanalization success during EST with an FPE of 45.7 % of mTICI0-occlusions and 44.7 % for mTICI1-occlusions (p = 0.709). This did not change when comparing ESTs, where only stent-retriever maneuvers under continuous aspiration were performed (FPE for mTICI 0: 100 (23.8 %) vs. mTICI 1: 40 (25.2 %), p = 0.745) or for thrombectomies, in which only contact aspiration maneuvers were performed (FPE for mTICI 0: 92 (21.8 %) vs. mTICI 1: 31 (19.5 %), p=0.571) – see table 2.

**Table 2.**
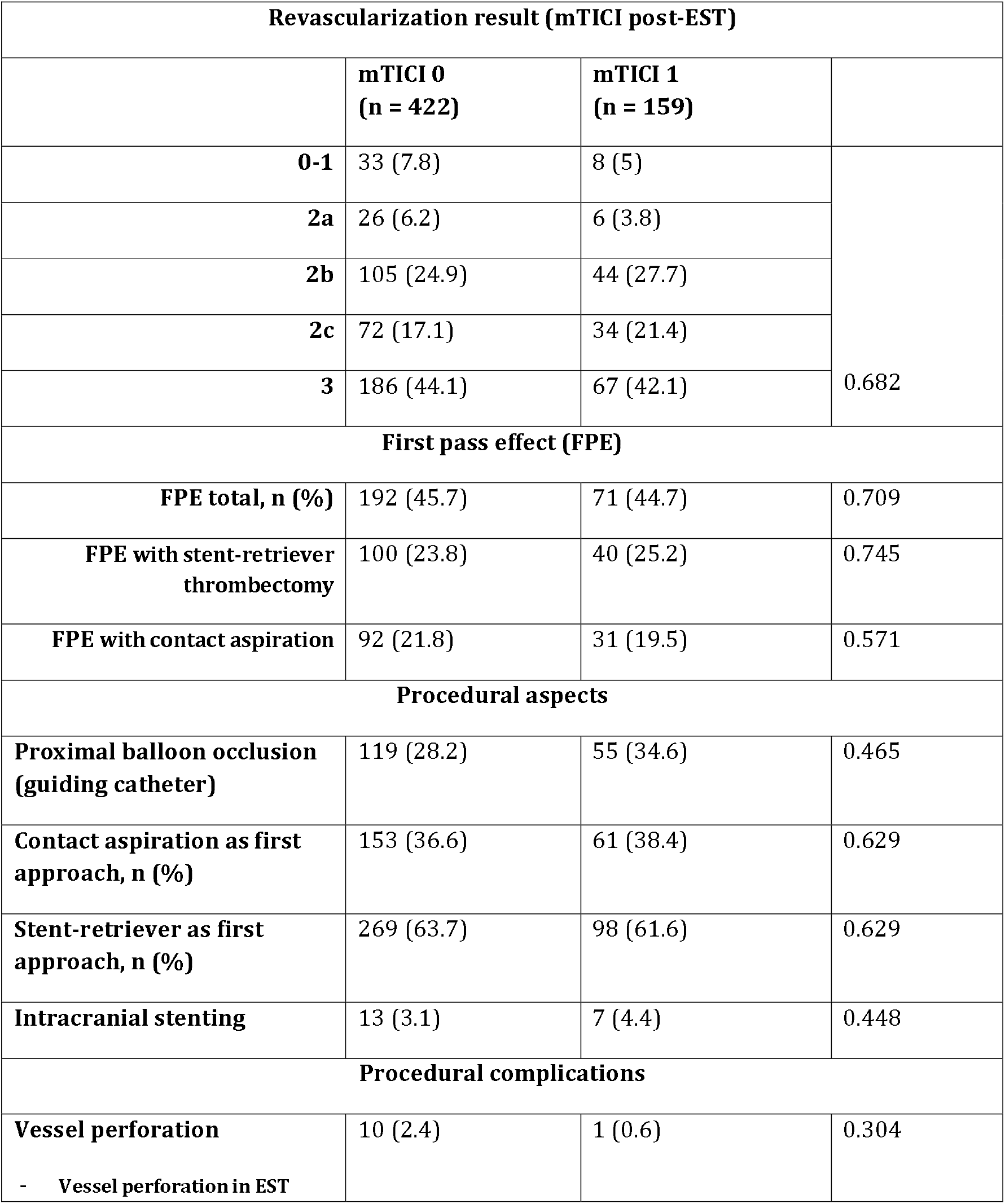
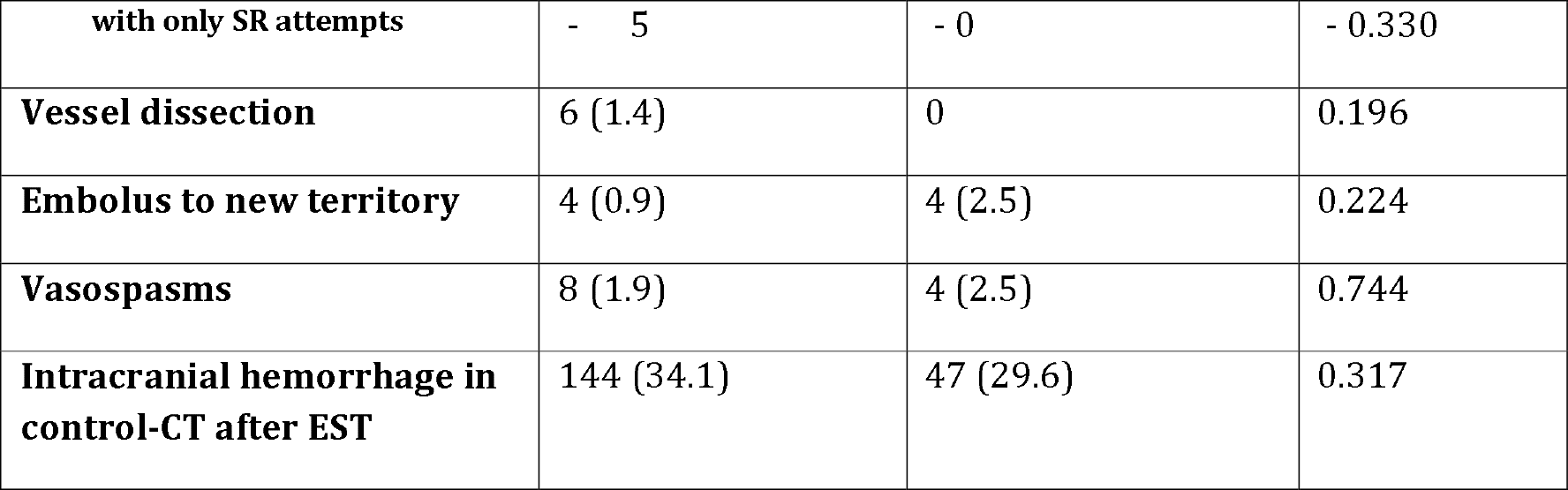
recanalization result and procedural complications of endovascular stroke treatment for complete (mTICI 0) and incomplete (mTICI 1) target vessel occlusions.

Concerning periprocedural complications, there were no statistically significant differences between study groups regarding vessel perforation, vessel dissection, thromboembolism to new territory during EST or postprocedural vasospasm. This did not change, when evaluating ESTs separately, in which only stent-retriever thrombectomy maneuvers were performed. The rate of intracranial hemorrhage in control imaging after EST was equally comparable – see table 2. While the mRS at discharge was lower for patients with incomplete occlusions (p = 0.014)), this difference did not persist and long-term clinical outcome measured as mRS 90 days after stroke onset did not differ between study groups – see table 2.

## Discussion

In this study, we found that an incomplete occlusion (mTICI 1) of an MCA M1-segment TVO eligible for EST is not associated with different recanalization results nor complication rate compared to complete target vessel M1-occlusion (mTICI 0). Also, when comparing the two major thrombectomy techniques – contact aspiration and stent-retriever thrombectomy – the FPE did not differ between study groups. Hence, we cannot conclude on the superiority of one technique over the other based on the initial mTICI Score. Compared to other studies describing TVO shapes, our study results are in line with Miranda et al., who did not find a differing recanalization rate for patients with meniscus sign in M1-occlusions^5^. In contrast, Garcia-Bermejo et al. reported a difference in the recanalization rate when comparing tapered and non-tapered occlusions with atherosclerotic disease underlying more often the tapered occlusions^9^. The complication rate did also not differ between the study groups, although the overall low incidence might have no statistical power since vessel perforation occurred four-fold more often in patients with a complete TVO (10 (2.4 %) vs. 1 (0.6 %), p = 0.304). The higher rate of vessel perforation in mTICI0-occlusions might be explained by the missing vessel contrast distal of the TVO in mTICI0-occlusions with a higher risk for vessel wall perforation in blind probing. However, there was also no difference between the study groups concerning the complication rates in stent-retriever thrombectomies.

Comparable to the influence of the hemispheric collateralization in intracranial large vessel occlusion (Tan-Score) on infarct growth of acute ischemic stroke and clinical outcome^10, 11^, the study’s mTICI 1 subgroup showed a lower NIHSS at hospital admission before EST and a lower mRS at discharge despite a comparable pre-stroke mRS and despite a higher patient age in patients with incomplete occlusions. However, the effect on clinical outcome did not persist over time and could not be found 90 days after stroke onset with a comparable mRS between study groups at this time point. Therefore, incomplete target vessel occlusions should be treated with the same urgency as complete target vessel occlusions. The higher ASPECTS after EST in the study group with incomplete occlusions may reflect a slower infarct growth. The difference in outcome and follow-up ASPECTS can be explained by the remaining blood flow through the TVO, which is the physiological correlate of mTICI Score 1 and persisting antegrade contrast perfusion.

Interestingly, patients with complete occlusions had been treated more often with intra-venous (i.v.) thrombolysis before EST. One might expect the contrary, i.e. i.v. thrombolysis as bridging lysis leading to incomplete occlusions in initial angiographic imaging of the TVO. On the other hand, we found more patients treated with antiplatelet medication and/or anticoagulants in the incomplete occlusion group, which means that many of these patients were not eligible for i.v. thrombolysis and, moreover, the previous antiplatelet or anticoagulant medication might also have an influence on thrombus composition and vessel occlusion properties.

Limitations of this study concern especially the single-center retrospective design. National or local specificities concerning EST’s procedural details might affect our study results. However, the large cohort reflecting many years of state of the art EST grant for a certain variety in individual approaches and technical details.

## Conclusion

Our study shows no difference concerning the recanalization success or the rate of complications between complete occlusions (mTICI0) and incomplete occlusions (mTICI1) of middle cerebral artery’s M1-segment target vessel occlusions. An initial effect on the clinical outcome at discharge (lower mRS for mTICI1 occlusions) did not persist at 90 days after stroke onset. Incomplete occlusions should therefore be treated with the same urgency as complete target vessel occlusions in the setting of acute ischemic stroke patients eligible for endovascular stroke treatment.

## Data Availability

All data produced in the present study are available upon reasonable request to the authors

